# Inhaled nitric oxide use in COVID19-induced hypoxemic respiratory failure

**DOI:** 10.1101/2021.08.19.21262314

**Authors:** Abhishek R. Giri, Siva Naga S. Yarrarapu, Nirmaljot Kaur, Alex Hochwald, Julie Crook, Scott Helgeson, Michael F. Harrison, Neal Patel, Pramod K. Guru, Philip Lowman, Pablo Moreno-Franco, Augustine Lee, Devang K. Sanghavi

## Abstract

**Introduction:** Nitric Oxide (NO) is an endogenous vasodilator that is synthesized by the vascular endothelium. Due to its vasodilatory effect and short half-life, the use of NO as an exogenous inhaled medication (iNO) to target the pulmonary vasculature, in conditions with increased pulmonary vascular resistance, has been studied.

The use of iNO in patients with ARDS secondary to COVID-19 has therapeutic importance in improving oxygenation. It also has potential anti-viral, anti-inflammatory, and anti-thrombotic properties.

Herein, we want to share our experience of use of iNO in hypoxemic respiratory failure secondary to COVID 19 pneumonia. We hypothesized that iNO may be beneficial at preventing intubation, decreasing invasive mechanical ventilation duration, and consequently improve outcomes including hospital mortality.

**Methods:** This is a descriptive hypothesis generating study of patients admitted for COVID-19 pneumonia who received iNO for hypoxemic respiratory failure, at a single tertiary care center. We collected information on patient demographics, co-morbidities, iNO treatment, need for intubation, arterial blood gas analysis, laboratory values, hospital length of stay, and mortality. Patients were divided into two groups based on the timing of iNO administration: group 1 - “pre-intubation” (i.e. iNO started at least 1 day prior to endotracheal intubation, if any) and group 2 - “post-intubation” (i.e. iNO started on the same day as or after endotracheal intubation and mechanical ventilation).

**Result:** A total of 45 (group 1, n=26 [57.8%] vs group2, n=19 [42.2%]) COVID 19 patients who had iNO use. The mean time from hospital admission to iNO administration(days) in group 1 was 2.1 (±1.8) vs 4.2 (±5.9) in group 2. The mean hospital length of stay from the beginning of iNO treatment until discharge or death was 18.3 vs 26.2 days, with 8 deaths (30.8%) vs 9 deaths (47.4%) in group 1 vs group 2, respectively.

**Discussion:** Our study is unable to demonstrate comparably outcomes benefit of iNO. Although there was a trend towards decreased need for invasive mechanical ventilation in group 1[Only 11 (42.3%) patients were intubated out of 26 who received iNO early after hospital admission (2.3 days)], no statistical significance could be achieved because of small sample size.

Our study demonstrated that iNO administration pre-intubation did not appear harmful and appears to be safe, complementary to HFNC, signalling the domain where systematic investigation is required to confirm or not the potential for iNO to improve patient outcomes in the management of COVID 19-induced hypoxemic respiratory failure.

**Conclusion:** This study showcases the potential benefit of early pre-intubation use of iNO in COVID patients with hypoxemic respiratory failure. This study could conclusively form the basis for a prospective trial and could have a tremendous impact in improving patient outcomes.

**Highlights:**

1. Inhaled nitric oxide can be used in the treatment COVID 19 induced hypoxemic respiratory failure.
2. Inhaled nitric oxide use can lower the burden on overwhelmed medical system.
3. Inhaled nitric oxide use may lower the need for intubation and subsequent invasive mechanical ventilation.

## Introduction

Nitric oxide (NO)is a potent vasodilator that is endogenously synthesized by the vascular endothelium. Due to its short half-life, inhaled NO(iNO) can provide vasodilation preferentially to ventilated areas improving ventilation-perfusion matching, in addition to potential benefits on pulmonary pressures and right heart afterload reduction.^1^

In critically ill patients, management of acute respiratory distress syndrome (ARDS) is predominantly supportive and without specific definitive treatment. The use of iNO in ARDS been relegated to “rescue” or “bridging” therapy for refractory hypoxemia. Although clinical trials have not been able to demonstrate a survival benefit of iNO in ARDS, iNO was universally applied late in the course of hypoxic respiratory failure, specifically, after they had required invasive mechanical ventilation. Early during the COVID-19 pandemic, facing scarce resources including mechanical ventilators and appropriately trained staff, our center initiated a treatment protocol of considering earlier use of iNO, pre-intubation, to manage refractory hypoxia and possibly delay need for endotracheal intubation or shorten duration of mechanical ventilator need. iNO has been shown to improve gas-exchange in a subset of patients affected by COVID-19 ARDS^2,3^, but it also has theoretical benefits in the specific context of COVID-19 pneumonia due to its anti-viral, anti-inflammatory, and anti-thrombotic properties.^1^

Hence, we aimed to explore whether pre-intubation administration of iNO resulted in any improved clinical outcomes among patients with COVID-19induced hypoxic respiratory failure. Herein, we want to share our experience of use of iNO in hypoxemic respiratory failure secondary to COVID 19 pneumonia. We hypothesized that iNO may be beneficial at preventing intubation, decreasing invasive mechanical ventilation duration, and consequently improve outcomes including hospital mortality.

## Methods

This is a descriptive hypothesis generating study of patients admitted during June 2020 to December 2020 with COVID-19 pneumonia who received iNO for hypoxemic respiratory failure, at a single tertiary care center. Criteria for iNO (5-20 ppm) addition to high flow nasal canula (HFNC) included patients with pure hypoxemia without increased work of breathing or respiratory acidosis, or if patient showed evidence of pulmonary hypertension (PH) and right ventricular (RV) failure. We collected information on patient demographics, co-morbidities, iNO treatment, need for intubation, arterial blood gas analysis, laboratory values, hospital length of stay, and mortality. Patients were divided into two groups based on the timing of iNO administration: group 1 - “pre-intubation” (i.e. iNO started at least 1 day prior to endotracheal intubation, if any) and group 2 - “post-intubation” (i.e. iNO started on the same day as or after endotracheal intubation and mechanical ventilation).Moreover, we compared two groups who required invasive endotracheal intubation (group 1) versus those who did not require intubation (group 2) as shown in table 3. We further analysed data based on survival at discharge and divided cohort into two groups: group 1 - alive and group 2 - deceased (refer table 4).

## Results

A total of 45 (group 1, n=26 [57.8%] vs group2, n=19 [42.2%]) COVID19 patients who had iNO use. Age and gender were similar between groups. (refer Table 1)

**Table 1:** Demographic variables by intubation relative to iNO ROX index – Predicts high flow nasal canula (HFNC) failure/need for intubation.

| Variable – Mean (SD) or<br>(Count, %) | iNO before<br>Intubation<br>(N=26) | iNO after<br>Intubation<br>(N=19) | Total (N=45) | p-value |
| --- | --- | --- | --- | --- |
| Sex (Female) | 9 (34.6%) | 7 (36.8%) | 16 (35.6%) | 1.000 |
| Age in years | 66.3 (10.7) | 63.7 (14.3) | 65.2 (12.2) | 0.512 |
| Race |  |  |  |  |
| • White | 17 (65.4%) | 12 (63.2%) | 29 (64.4%) | 0.743 |
| • African American | 5 (19.2%) | 4 (21.1%) | 9 (20.0%) |  |
| • Asian | 2 (7.7%) | 2 (10.5%) | 4 (8.9%) |  |
| • Hispanic | 2 (7.7%) | 0 (0.0%) | 2 (4.4%) |  |
| • Pacific Islander | 0 (0.0%) | 1 (5.3%) | 1 (2.2%) |  |
| SOFA score within 24 hours of hospitalization | 2.8 (1.5) | 6.3 (2.9) | 4.3 (2.8) | <0.001 |
| ROX index within 24 hours of hospitalization | 5.7 (3.5) | 4.0 (1.9) | 5.0 (3.0) | 0.063 |
| Smoking status<sup>1</sup><br>• Current smoker<br>• Former smoker<br>• Never smoked | 1 (3.8%),<br>7 (26.9%),<br>18 (69.2%) | 1 (5.3%),<br>2 (10.5%),<br>15 (78.9%) | 2 (4.4%),<br>9 (20.0%),<br>33 (73.3%) | 0.384 |
| APACHE II score at the time of iNO administration | 11.7 (4.1) | 22.4 (7.5) | 16.2 (7.8) | <0.001 |
| Time between admission to iNO treatment initiation (days) | 2.1 (1.8) | 4.2 (5.9) | 3.0 (4.2) | 0.815 |

The mean time from hospital admission to iNO administration(days) in group 1 was 2.1 (±1.8) vs 4.2 (±5.9) in group 2. Severity of illness at admission as indicated by the mean Sequential Organ Failure Assessment (SOFA) score which predicts ICU mortality based on lab results and clinical data, and the APACHE-II scores were worse in group 2 than in group1. The mean ventilator-free days (VFD) was 4.3 in group 1 as opposed to 6.4 in group 2, although this was not statistically different. The mean duration of mechanical ventilation was 12.6 vs 19.1 days in group 1 vs group 2, respectively. The mean hospital length of stay from the beginning of iNO treatment until discharge or death was 18.3 vs 26.2 days, with 8 deaths (30.8%) vs 9 deaths (47.4%) in group 1 vs group 2, respectively (refer Table 2.).

**Table 2:** Outcome factors by intubation relative to iNO Ventilator-free days = 28 – x, if successfully liberated from ventilation x days after initiation. VFDs = 0 if subject dies within 28 days of mechanical ventilation.

| Variable - Mean (SD) or<br>(Count, %) | iNO before<br>Intubation<br>(N=26) | iNO after<br>intubation (N=19) | Total (N=45) | p-value |
| --- | --- | --- | --- | --- |
| Patient intubated at any<br>time (Yes) | 11 (42.3%) | 19 (100.0%) | 30 (66.7%) | <0.001 |
| Total duration of invasive<br>mechanical ventilation<br>(days) | 12.6 (9.7) | 19.1 (19.0) | 16.7 (16.3) | 0.605 |
| Ventilator- free days | 4.3 (9.6) | 6.4 (9.9) | 5.6 (9.7) | 0.562 |
| Total length of hospital<br>stays (days) | 20.4 (13.3) | 30.4 (24.6) | 24.6 (19.3) | 0.29 |
| Length of hospital stay<br>after iNO treatment<br>initiation (days) | 18.3 (12.7) | 26.2 (23.6) | 21.6 (18.3) | 0.427 |
| Vital status at discharge<br>(deceased) | 8 (30.8%) | 9 (47.4%) | 17 (37.8%) | 0.353 |

In table 3, we compared two groups, group 1(required invasive endotracheal intubation) and group 2 (did not require intubation). High SOFA score at the time of admission associated with increased need for intubation during hospitalization.

**Table 3:** Demographic variables and outcome factors by intubation status

| Variable - Mean (SD) or<br>(Count, %) | No intubation<br>at any time<br>(N=15) | Patient intubated<br>(N=30) | Total (N=45) | p-value |
| --- | --- | --- | --- | --- |
| Sex (Female) | 6 (40.0%) | 10 (33.3%) | 6 (40.0%) | 0.746 |
| Age in years | 64.1 (11.2) | 65.8 (12.9) | 65.2 (12.2) | 0.736 |
| Race |  |  |  | 0.406 |
| • White | 3 (20.0%) | 6 (20.0%) | 9 (20.0%) |  |
| • African American | 1 (6.7%) | 3 (10.0%) | 4 (8.9%) |  |
| • Asian | 2 (13.3%) | 0 (0.0%) | 2 (4.4%) |  |
|  | 0 (0.0%) | 1 (3.3%) | 1 (2.2%) |  |
| • Hispanic | 9 (60.0%) | 20 (66.7%) | 29 (64.4%) |  |
| • Pacific Islander |  |  |  |  |
| SOFA score within 24<br>hours of hospitalization | 2.6 (1.4) | 5.1 (3.0) | 4.3 (2.8) | 0.003 |
| APACHE II score at the<br>time of iNO administration | 9.9 (3.2) | 19.3 (7.6) | 16.2 (7.8) | <0.001 |
| Time between admission to iNO treatment beginning (days) | 2.3 (2.2) | 3.3 (4.9) | 3.0 (4.2) | 0.873 |
| Total duration of invasive mechanical ventilation (days) |  | 16.7 (16.3) | 16.7 (16.3) |  |
| Ventilation free days |  | 5.6 (9.7) | 5.6 (9.7) |  |
| Total length of hospital stays (days) | 16.5 (12.1) | 28.7 (21.1) | 24.6 (19.3) | 0.030 |
| Length of hospital stay after iNO treatment began (days) | 14.2 (10.7) | 25.4 (20.3) | 21.6 (18.3) | 0.031 |
| Patient's vital status at discharge (Deceased) | 0 (0.0%) | 17 (56.7%) | 17 (37.8%) | <0.001 |

In table 4, we compared two groups by their survival at discharge, group 1(alive) vs group 2(deceased).In group 1, 13 (46.4%) patients were intubated in comparison to 17 (100%) patients in group 2.Even though numerical values showed correlation to improved survival with decreased time between admission to iNO administration, statistically it is not significant.

**Table 4:** Demographic variables and outcome factors by death status.

| Variable - Mean (SD) or<br>(Count, %) | Alive at<br>discharge<br>(N=28) | Deceased (N=17) | Total (N=45) | p-value |
| --- | --- | --- | --- | --- |
| Sex (Female) | 12 (42.9%) | 4 (23.5%) | 16 (35.6%) | 0.219 |
| Age in years | 61.2 (12.5) | 71.8 (8.7) | 65.2 (12.2) | 0.006 |
| Race |  |  |  | 0.097 |
| • White | 14 (50.0%), | 15 (88.2%), | 29 (64.4%), |  |
| • African American | 8 (28.6%), | 1 (5.9%), | 9 (20.0%), |  |
| • Asian | 3 (10.7%), | 1 (5.9%), | 4 (8.9%), |  |
| • Hispanic | 2 (7.1%), | 0 (0.0%), | 2 (4.4%), |  |
| • Pacific Islander | 1 (3.6%) | 0 (0.0%) | 1 (2.2%) |  |
| SOFA score within 24<br>hours of hospitalization | 3.8 (2.6) | 5.1 (3.0) | 4.3 (2.8) | 0.114 |
| APACHE II score at the<br>time of iNO administration | 13.5 (6.0) | 20.5 (8.7) | 16.2 (7.8) | 0.006 |
| Time between admission<br>to iNO treatment<br>beginning (days) | 2.9 (4.3) | 3.2 (4.0) | 3.0 (4.2) | 0.784 |
| Intubation Status<br>(Ventilated) | 13 (46.4%) | 17 (100.0%) | 30 (66.7%) | <0.001 |
| Total duration of invasive<br>mechanical ventilation<br>(days) | 21.3 (19.7) | 13.2 (12.7) | 16.7 (16.3) | 0.63 |
| Length of hospital stay<br>after iNO treatment began<br>(days) | 25.1 (20.6) | 15.9 (12.4) | 21.6 (18.3) | 0.246 |
| Total length of hospital<br>stays (days) | 28.0 (21.4) | 19.1 (14.2) | 24.6 (19.3) | 0.302 |

Kaplan Meier curve below estimates that iNO administration delays intubation by about 7 days. The log-rank statistic is p= 0.0447.

**Figure 1.**
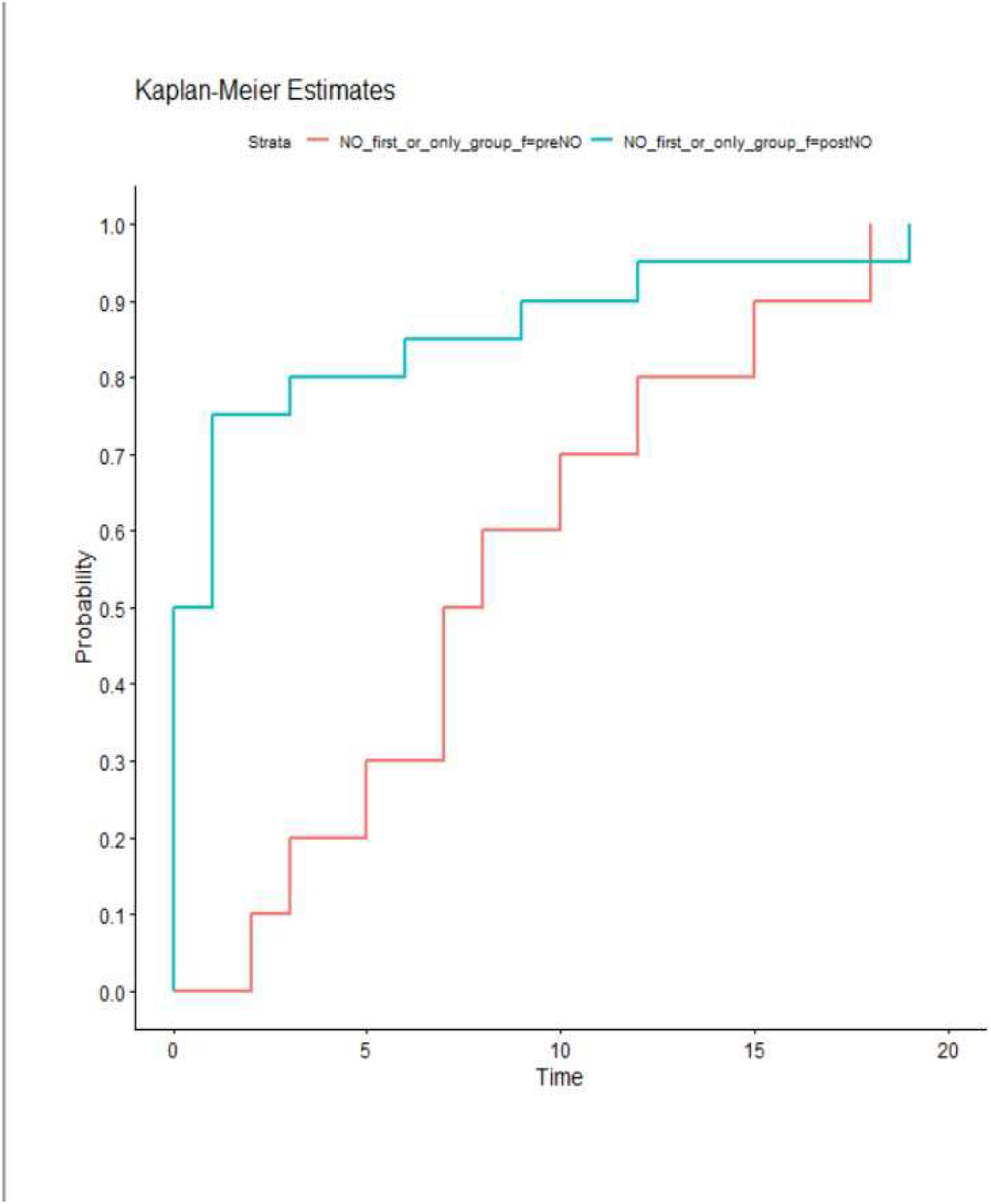
Kaplan Meier curve showing effects of iNO use on intubation.

## Discussion

In this study, we have summarized the available data and underlined the hypoxemic respiratory failure scenario wherein iNO can be used. Although our study was unable to demonstrate comparably the outcomes of iNO use in either group, across the three different analyses of COVID-19 patients, there was a trend of decreased need for invasive mechanical ventilation in group 1. In group 1 only 11 (42.3 %) patients were intubated out of 26 who received iNO early on after hospital admission (2.3 days), [no statistical significance could be achieved because of small sample size]. High mortality (56.7%) was observed in intubated patients (group 2, table 3) irrespective of iNO administration timing (pre-intubation vs post-intubation). Better SOFA (2.6) and APACHE (9.9) scores were associated with in group 1 (refer table 3) resulting in no need for mechanical ventilation. Overall summarising potentially favourable outcomes (decreased mortality) if nitric oxide was administered earlier in patients with less severity.

In recent years, high flow nasal cannula (HFNC) use has been increased prior to intubation in management of hypoxemic respiratory failure. High flow nasal cannula delivers humidified oxygen and it washes out dead space in hypoxemic respiratory failure. It increases CO2 clearance, decreases respiratory drive, and facilitate effortless breathing with decreased respiratory rate.^4^ In addition to this, Abou-Arab et al. demonstrated an improvement of PaO2/FiO2 ratio with iNO use in COVID-19 patients with severe pneumonia.^2^ Improved arterial oxygenation was also shown by Lotz et al. wherein increased partial pressure of oxygen was seen subsequent to iNO administration in COVID-induced ARDS.^3^ Nitric oxide inhibits COVID 19 virus replication and its attachment to lung ACE2 receptors. Inhaled NO has anti-inflammatory properties reducing inflammatory mediators induced lung injuries. It improves ventilation/perfusion match, decreases pulmonary vascular resistance as well as transfusion of fluid into the alveolar spaces.^5^ Although no response was demonstrated with iNO in a few studies, design of these studies precluded any formal impression of improved outcome with iNO use.^6,7^ These studies investigated the use of iNO in COVID-19 with mixed results varying from positive outcomes to a lack of any effect, but did not address the effects of iNO use on intubation and subsequent invasive mechanical ventilator support. Our study demonstrated that iNO administration pre-intubation was not harmful and appears to be safe, complementary to HFNC, signalling the domain where systematic investigation is required to confirm the potential for iNO to improve patient outcomes in the management of COVID 19-induced hypoxemic respiratory failure.

### Study Limitations

This is retrospective study with small sample size; hence desired results were not statistically significant. The effects of iNO were not charted in terms of patient outcomes. We collected SOFA score on admission. Therefore, the severity of patient condition may have deteriorated or improved after 24 hrs. Potential confounders include the novel nature of this virus with rapidly changing standards of medical care, which are not included in this study.

## Conclusion

This study showcases the potential benefit of early pre-intubation use of iNO in COVID-19 patients with hypoxemic respiratory failure. This study could conclusively form the basis for a prospective trial and could have a tremendous impact in improving patient outcomes. Thus, more studies are needed in future to evaluate the effects of iNO use in hypoxemic respiratory failure.

## Data Availability

We would like to declare that all data referred to in the manuscript is available with us.

## Abbreviations

iNO: (Inhaled nitric oxide)
ICU: (Intensive care unit)
SOFA: (Sequential Organ Failure Assessment)
SD: (Standard Deviation)

